# Regional variations in primary percutaneous coronary intervention for acute myocardial infarction patients: A trajectory analysis using the national claims database in Japan

**DOI:** 10.1101/2022.10.12.22280985

**Authors:** Hisashi Itoshima, Jung-ho Shin, Noriko Sasaki, Etsu Goto, Susumu Kunisawa, Yuichi Imanaka

## Abstract

v.

**BACKGROUND:** Previous studies have demonstrated geographical disparities regarding the quality of care for acute myocardial infarction (AMI). The aim of this study was two-folds: first, to calculate the proportion of patients with AMI who received primary percutaneous coronary interventions (pPCIs) by secondary medical areas (SMAs), which provide general inpatient care, as a quality indicator (QI) of the process of AMI practice. Second, to identify patterns in their trajectories and to investigate the factors related to regional differences in their trajectories.

**METHODS:** We included patients hospitalized with AMI between April 2014 and March 2020 from the national health insurance claims database in Japan and calculated the proportion of pPCIs across 335 SMAs and fiscal years. Using these proportions, we conducted group-based trajectory modeling to identify groups that shared similar trajectories of the proportions. In addition, we investigated area-level factors that were associated with the different trajectories.

**RESULTS:** The median (interquartile range) proportions of pPCIs by SMAs were 63.5% (52.9% to 70.5%) in FY 2014 and 69.6% (63.3% to 74.2%) in FY 2020. Four groups, named low to low (LL; n =, low to middle (LM; n = 16), middle to middle (MM; n = 68), and high to high (HH; n = 208), were identified from our trajectory analysis. The HH and MM groups had higher population densities and higher numbers of physicians and cardiologists per capita than the LL and LM groups. The LL and LM groups had similar numbers of physicians per capita, but the number of cardiologists per capita in the LM group increased over the years of the study compared with the LL group.

**CONCLUSION:** The trajectory of the proportion of PCIs for AMI patients identified groups of SMAs. Among the four groups, the LM group showed an increasing trend in the proportions of pPCIs, whereas the three other groups showed relatively stable trends.

**Summary boxes:** *What is already known on this topic:*

- The quality of health care for acute myocardial infarction (AMI) patients varies across regions.
- Door-to-balloon time is associated with patient outcomes, and one of the most easily measurable indicators is the provision of rapid treatment for AMI patients.

*What this study adds:*

- The study demonstrated that there were not only cross-sectional regional differences in the proportion of PCIs for AMI patients but also differences in these trajectories.
- The cross-sectional regional differences in the proportion of pPCIs were maintained in many SMAs, but there were a few SMAs that increased yearly.

## INTRODUCTION

Health equity is one of the most important concerns in public health. The World Health Organization states several health equity policy action areas and indicators to ensure access to health services of equally good quality [1]. According to Donabedian’s model, the quality of health care is divided into three components: structure, process, and outcome [2]. Process indicators refer to what health care providers do for patients. These include adherence to standard treatments advised in clinical practice guidelines, such as door-to-balloon time for patients requiring primary percutaneous coronary intervention (pPCI) and the use of antiplatelet drugs at arrival for patients with acute myocardial infarction (AMI) patients [3].

Previous studies have demonstrated geographical variation regarding the quality of care for AMI and other acute diseases [3, 4, 5, 6, 7, 8]. For example, the use of beta-blockers after AMI is recommended, but the proportion of beta-blocker usage was low and geographical variation was observed [4]. PCI is a favorable treatment strategy for patients with ST-segment elevation myocardial infarction (STEMI). The American College of Cardiology Foundation/American Heart Association guidelines 2013 for STEMI recommends that primary PCI should be conducted within 90 min of arrival at the hospital [9]. The Japanese circulation society 2018 Guideline on Diagnosis and Treatment of Acute Coronary Syndrome recommends a door-to-device time shorter than 90 min as the minimum acceptable time, and total ischemic time should be as short as possible [10]. The time until PCI (door-to-balloon time) is related to patient outcomes, and the provision of rapid treatment is one of the easiest quality of care indices to measure for AMI patients [11]. Although, it can be inferred that it is preferable for the status of pPCI to be fair across geographic regions, based on previous papers mentioned above. Not all AMI patients are eligible for pPCI, but it was performed in 72.5% of patients (83,658/115,407) in Japan [12]; however, there have been few studies on the geographic variation in pPCI for AMI patients.

The aim of this study was two-folds: first, to calculate the proportion of patients with AMI who received pPCI by secondary medical areas (SMAs), which provide general inpatient care, as a quality indicator (QI) of the process of AMI practice. Second, to identify patterns in their trajectories and investigate the factors related to regional differences in their trajectories.

## METHODS

### Data source

We conducted this study using the National Database of Health Insurance Claims and Specific Health Checkups of Japan (NDB) administrated by the Japanese Ministry of Health, Labor and Welfare (MHLW) since 2008. Japan had a universal health coverage system since the 1960s [13], and all health insurance claims data have been collected by MHLW since 2009. Therefore, this large database includes the health insurance claims data of nearly all (≥90% in 2014) medical practices such as procedures, examinations, drug prescriptions, etc. [14, 15]. Since 2012, more than 1.6 billion claim records have been registered in the NDB annually, and MHLW has given permission to use this data for research and health policy-making [14, 15, 16]. The NDB contains age, sex, diagnoses, date of consultation, date of admission, procedure, medication, and health checkup data.

### Study population

We included AMI patients who were hospitalized between April 1, 2014, and March 30, 2020, from the NDB. The definition of AMI patients was that they had been diagnosed with AMI (ICD-10 code: I21x) and received blood tests for cardiac biomarkers (troponin I, etc.) or an echocardiogram or ultrasonic echocardiography or coronary angiography or chest X-ray based on the guidelines in Japan [17].

### Regional units: Secondary medical areas in Japan

Japan consists of 47 prefectures and 1718 municipalities. The medical provision system is based on the medical care plan, which is made by prefectural governments [18, 19]. The Japanese medical system has a three-level hierarchy: primary medical areas, secondary medical areas, and tertiary medical areas. Primary medical areas provide primary care at a municipal level, secondary medical areas (SMAs) provide general inpatient care at several municipal levels, and tertiary medical areas are responsible for tertiary medical care at a prefectural level. SMAs have been restructured since 2014, with 335 units in June 2022. The data for SMAs before the reorganization were adjusted to those after the reorganization, and the data for 335 SMAs were used for this analysis [18, 19].

### Outcomes

We calculated the proportion of pPCIs (within one day after admission) for AMI patients in each SMA (adjusted by a shrinkage estimate). We supposed that the number of pPCIs based on actual measurements was unstable due to wide variation among the AMI patients of different SMAs. To compensate for this assumption, a shrinkage estimation was introduced by taking into account the population of the SMA and converging it to the average of each prefecture. This is the same method used to calculate the English indices of deprivation (the IoD) [20].

We calculated the shrinkage estimate for pPCI for AMI patients based on the methods used for the IoD [20].

The logit *SMA*_*j*_ for an event occurrence *r*_*j*_ for a population *n*_*j*_in a *j*th *SMA* is as follows:

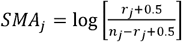

The estimated standard error *S*_*j*_ is

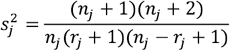

The logit *Pref* in a prefecture to which *j*th *SMA* belongs is calculated using a population *n* and event *r* in a prefecture

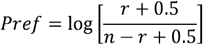

The “shrinkage” logit 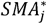 in a *j*th *SMA* with low event occurrence is a weighted average.

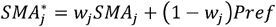

*W*_*j*_ is a weight given to an SMA j where the event occurrence is low, and (1−*w*_*j*_) is a weight given to a prefecture to which it belongs *w*_*j*_ is calculated using standard error *s*_*j*_ and variance *t*^2^.

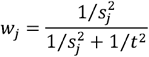

Where *t*^*2*^ is the inter-SMAs variance for the k lower-layer SMAs in a prefecture, calculated as:

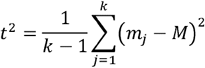

### Statistical analysis

### Trajectory analysis

In recent studies, more attention has been given to the analysis of longitudinal data, typically using multilevel models. These model results estimate the average trajectory of a population and the variation of individual-level trajectories around this average [21, 22]. However, the identification of several typical trajectories, rather than just one average trajectory, and more detailed modeling of these individual variations, may be a more useful approach to exploring common subgroups. [21, 22, 23, 24, 25] These methods can be classified into:

- Growth mixture models
- Latent class growth analysis (as group-based trajectory models)
- Longitudinal latent class analysis

Several published articles examine the association between socioeconomic factors at an individual level and regional differences using trajectory model analysis [26, 27, 28]. We conducted a group-based trajectory model analysis for separating clusters according to their similarity in terms of their time courses of the proportion of pPCIs in each SMA using the latrend package (version 1.4.1) for R [29]. The optimal number of clusters was decided by BIC, and the mean absolute error of the fitted trajectories was weighted by cluster assignment probability (WMAE) [29]. The fitted function was selected as linear or quadratic, spline based on the trajectory of the proportion of pPCIs. A two-sided P-value <0.05 was considered statistically significant, and all analyses were conducted using R 4.1.2 (R Foundation for Statistical Computing, Vienna, Austria).

### Post-hoc analysis

To examine geographic and socioeconomic, health care factors associated with regional differences among groups of varied pPCI trajectories, the following parameters were used as area-level factors. Geographic and socioeconomic factors included the population, the proportion of the population over 65 years of age, population density, unemployment rate, and per capita taxable income. Health care factors included: the number of total medical doctors per 100,000 population, cardiologists per 100,000 population, dentists per 100,000 population, pharmacists per 100,000 population, general hospitals per 100,000 population, emergency hospitals per 100,000 population and general hospital beds per 100,000 population [19, 28]. These variables were collected from the population census in 2015 in Japan and the 2016 survey of physicians, dentists, and pharmacists, and the 2016 survey of medical institutions from the MHWL [30, 31, 32]. Regarding healthcare-related factors, the trends in the number of medical doctors, cardiologists, general hospitals, and emergency hospitals between 2014 and 2018 were also obtained from the 2014 and 2018 surveys of physician, dentists, and pharmacists, and the 2014 and 2018 surveys of medical institutions from the MHWL [33, 34, 35, 36]. We displayed these group characteristic variables using a median in each group. The definition of a general hospitals included all hospitals except psychiatric hospitals, and the definition of emergency hospitals included hospitals that provided any emergency medical services at the primary, secondary, or tertiary level [33, 34].

### Ethical consideration

The present study was approved by the ethics committee of Kyoto University (approval number: R2215). Informed consent was not required because the data was anonymous, in accordance with the Ethical Guidelines for Medical and Health Research Involving Human Subjects, as stipulated by the Japanese Government. The authors of the manuscript have no conflicts of interest.

## RESULTS

Overall, 469,289 cases with an AMI diagnosis were included in this study, and during the study period, there were 322,779 pPCI procedures for AMI patients among the 355 SMAs. The time trend in the shrinkage estimated proportion of pPCIs for AMI patients showed large variation in each SMA (Figure1).

In Figure 1, it can be seen that linear and quadratic terms did not fit well. Therefore, the spline function was used. We also calculated the optimal number of trajectories between 1 to 5 by BIC and WMAE, then determined that a suitable number of trajectories was 4, based on BIC and WMAE (Figure 2). Model adequacy was as follows: WRSS was 17.593, WMAE was 0.062 and BIC was -4398.594. The 335 SMAs were classified into four groups: group A had 208 SMAs (62.0%), group B had 68 SMAs (20.0%), group C had 16 SMAs (5.0%), and group D had 43 SMAs (13.0%). Our model also classified similar trajectories well (Figure 3). Each group was named based on its trajectory: group A was the high to high group (HH group), group B was the middle to middle group (MM group), group C was the low to middle group (LM group), and group D was the low to low group (LL group).

**Figure 1.**
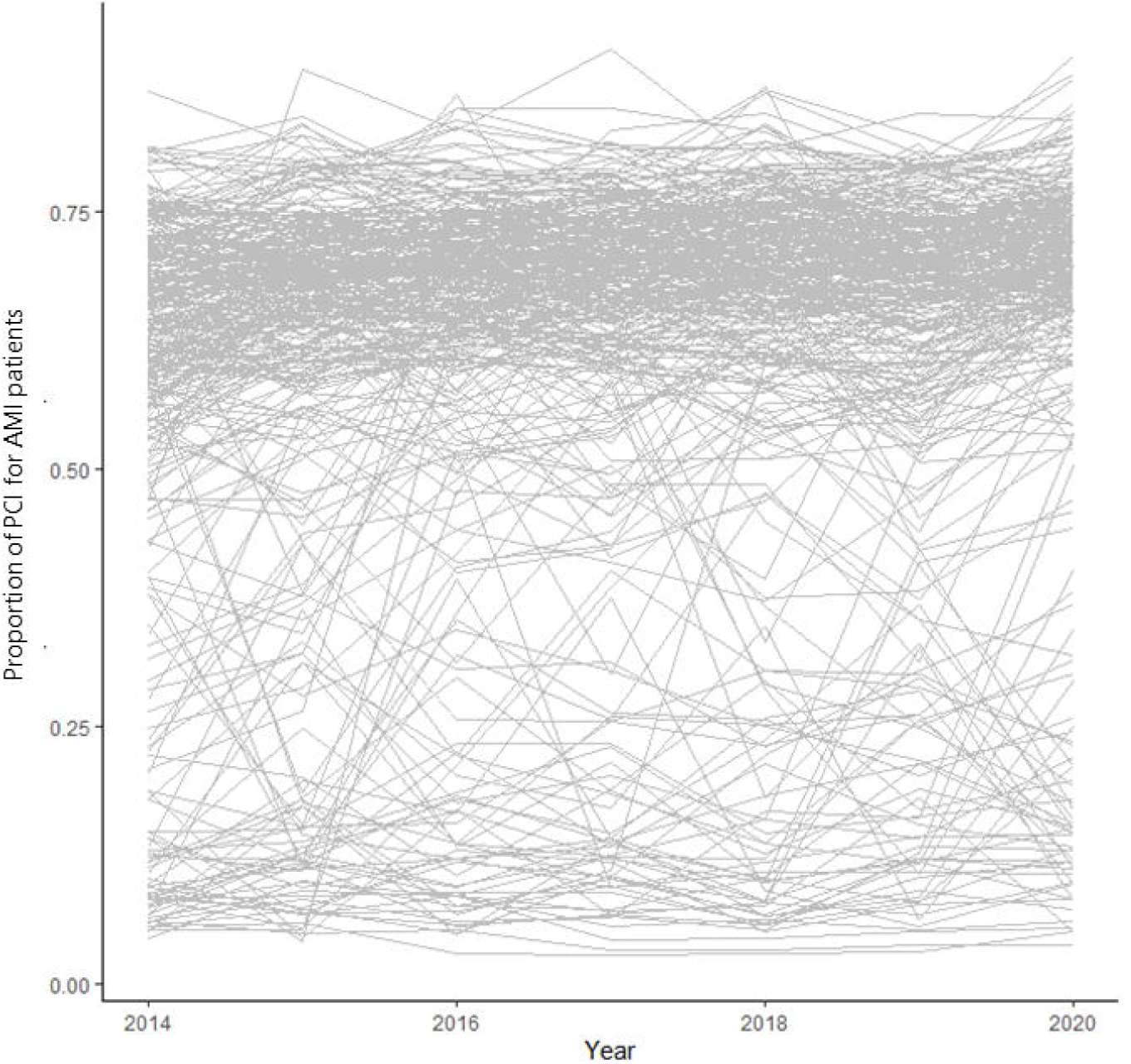
Spaghetti plot of the proportion of urgent primary percutaneous coronary intervention (pPCIs) for AMI patients by each secondary medical areas

**Figure 2.**
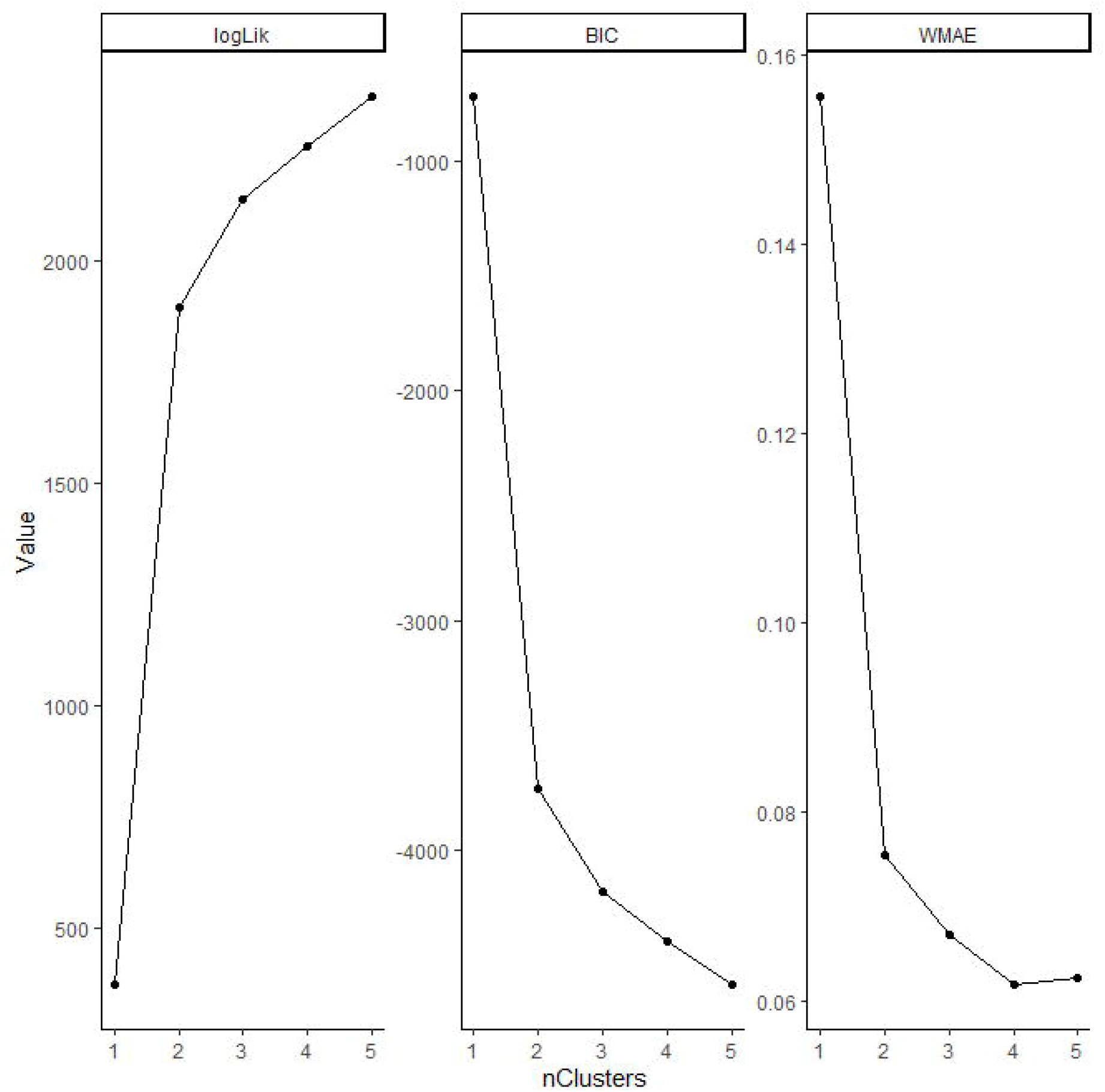
Results of group-based trajectory modeling (spline function) by the number of groups

**Figure 3.**
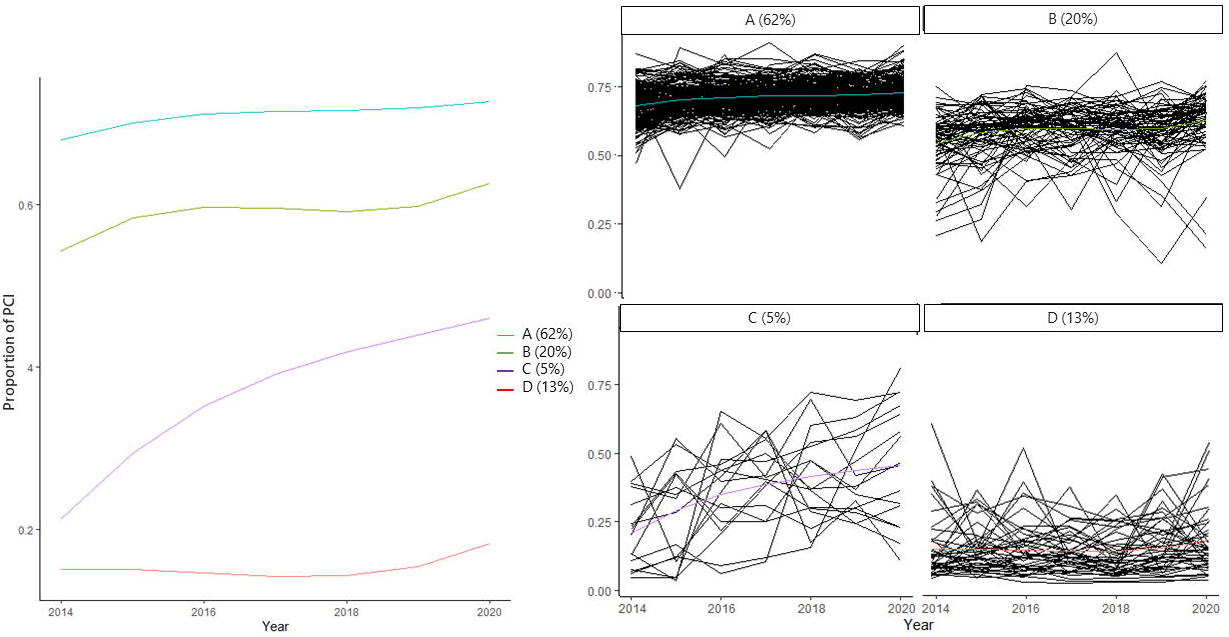
The trends in each group of trajectories

The characteristics of each group, such as its population, population density, unemployment rate, and the number of physicians per 100,000 capita, are reported in Table 1. Groups A and B had a greater population, population density, and medical professions than groups C and D. Supplementary figures demonstrate the distribution of the population, the proportion of the population over 65 years of age, the total number of medical doctors per 100,000 population, cardiologists per 100,000 population, general hospitals per 100,000 population, and general hospital beds per 100,000 population in each group (eFigures 1, 2, 3, 4, 5, and 6).

**Table 1.** The characteristics of each group by group-based trajectory modeling

| Group (n* = 335) | Group A (n = 208) | Group B (n = 68) | Group C (n = 16) | Group D (n = 43) |
| --- | --- | --- | --- | --- |
|  | High to high group | Middle to middle group | Low to middle group | Low to low group |
| **Geographic and socioeconomic factors** |  |  |  |  |
| Population, median | 348,697 | 158,241 | 78,331 | 66,438 |
| 1stQR, 3rdQR | 182,155 to 646,332 | 96,065 to 268,407 | 38,860 to 127,815 | 52,517 to 91,084 |
| Proportion of 65 years older | 0.256 | 0.265 | 0.316 | 0.334 |
| Population density | 14.941 | 8.945 | 3.961 | 2.405 |
| Unemployment rate | 0.043 | 0.042 | 0.045 | 0.039 |
| Per capita taxable income (1,000 yen) | 3,019 | 3,237 | 2,374 | 2,222 |
| **Health Care Factors\*\*** |  |  |  |  |
| Medical doctor | 190.4744 | 193.345 | 140.786 | 143.858 |
| Cardiologists | 7.773 | 7.807 | 3.835 | 3.734 |
| Dentists | 64.618 | 90.670 | 63.192 | 56.964 |
| Pharmacists | 187.902 | 256.084 | 167.814 | 148.437 |
| General hospitals | 5.456 | 8.456 | 6.570 | 9.008 |
| Emergency hospitals | 3.078 | 4.052 | 4.095 | 5.920 |
| General hospital beds | 561.009 | 678.675 | 592.142 | 717.986 |

\*n is the number of secondary medical areas
\*\*All variables per 100,000 population
† All variables were median in each group

**Figure 4.**
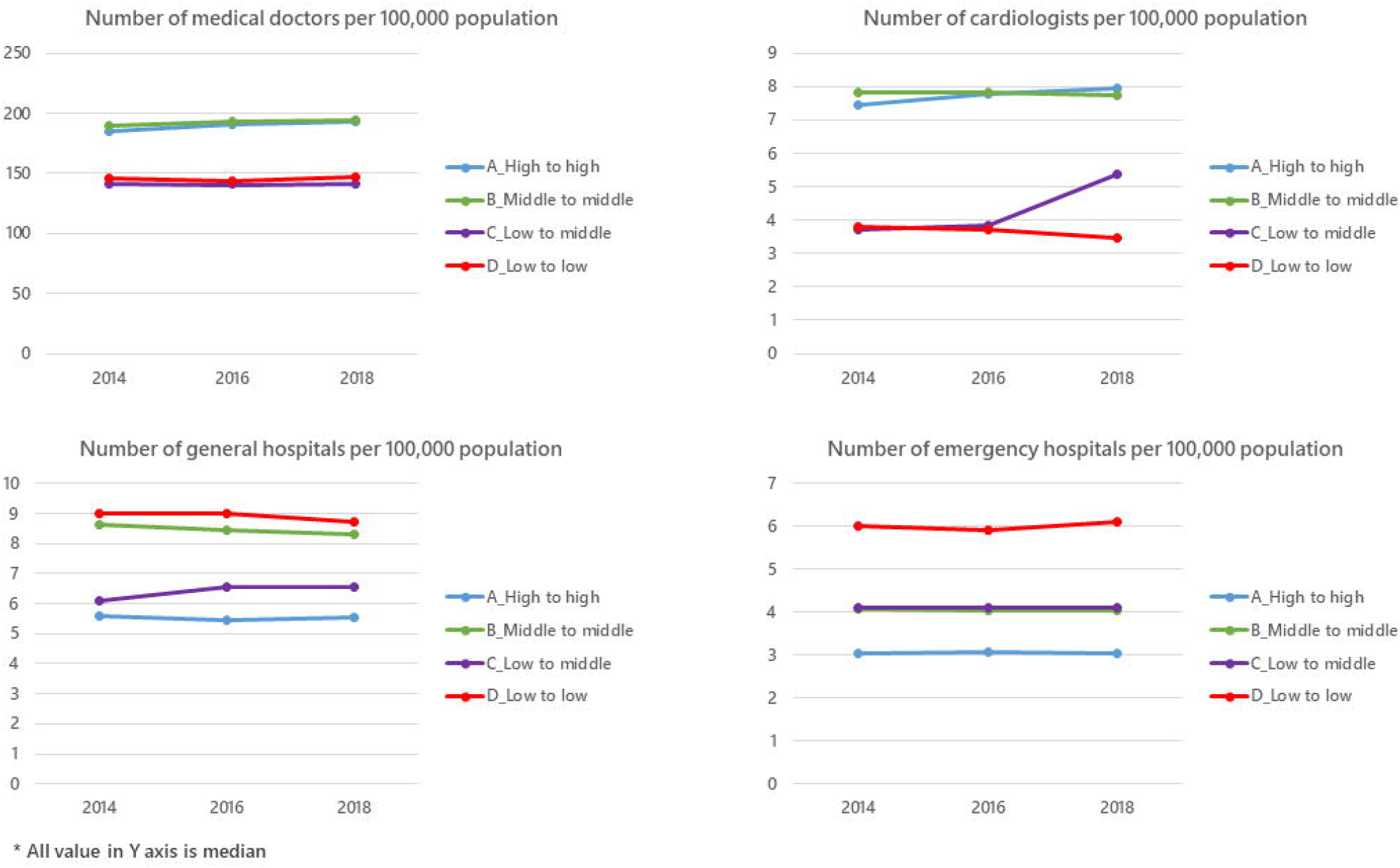
The trends in the number of medical doctors, cardiologists, general hospitals, and emergency hospitals between 2014 and 2018

The trends in the number of physicians, cardiologists, general hospitals, and emergency hospitals between 2014 and 2018 are shown in Figure 4. Each group had no change regarding the number of hospitals; however, cardiologists increased only in the LM group (4 to 6 per 100,000 population).

## DISCUSSION

We found geographical differences in the proportion of pPCIs for AMI patients and their trends over time in Japanese SMAs between 2014 and 2020. Furthermore, our trajectory modeling analysis displayed four groups of SMAs using group-based trajectory analysis of the trends in the proportion of pPCIs for AMI patients and assessed whether area-level factors related to this distribution. The groups were classified into those with a medium to high proportion of pPCIs over time (the HH and the MM groups), those with a low proportion over time (the LL group), and those initially with a low but gradually increasing proportion over time (the LM group). In terms of geographic, socioeconomic, and healthcare-related factors, the HH and MM groups were those with large populations and abundant healthcare resources, while the LL and LM groups were those with small populations and few healthcare resources. Compared to the LL group, the total number of physicians remained the same in the LM group, but the number of cardiologists increased over time in the LM group.

### Strengths and weaknesses of the study

Regional disparities in the proportion of AMI patients who received PCIs were assessed in this study form the NDB, which contains all medical services covered by the National Health Insurance Scheme, including PCIs. Therefore, our study could analyze national-level regional differences in this topic using complete longitudinal data. The time to balloon is a quality indicator of PCI; therefore, it might be desirable to complete PCI procedures in a region close to the patient’s home [37, 38]. The regional difference in the proportion of AMI patients who received a pPCI indicated regional disparities in the quality of health care for AMI patients. There were several studies discussing quality indicators related to AMI, including PCI and regional differences [4, 5, 6, 7, 37, 38, 39]. Several cross-sectional studies have discussed socioeconomic factors, health care resources, and healthcare performance regarding AMI practice [40, 41, 42]. This study examined changes in quality indicators over seven years and identified a group that initially had a low proportion of pPCIs, which improved over time (the LM group).

Also, we found a difference in the trajectory of the proportion of pPCIs among lower groups at the beginning of the study (the LL group and LM group), which might be related to the number of cardiologists increasing between 2014 and 2018 in the LM group. Studies examining the density of specialists and outcomes in a surgical field have shown that colorectal cancer mortality is higher in areas with fewer colorectal specialists and prostate cancer mortality is lower in areas with radiation oncologists than in areas without [43, 44]. However, few studies have examined the relationship between an increase in cardiologists and improvements in the quality of health care regarding AMI practice. The reason behind the increasing number of cardiologists in the LM group was unclear from our data. One possible explanation of this phenomenon might occur the centralization of cardiologists in the LM group. Park et al. indicated that the centralization of cardiologists at a hospital level is associated with lower in-hospital mortality; therefore, the centralization of cardiologists might occur in this group [45]. This study had several limitations. First, our study only evaluated the proportion of pPCI procedures in SMAs, which is a process indicator for pPCI, but could not evaluate other process indicators such as the administration of acetylsalicylic acid before pPCI or, the proportion of PCIs with GFR documentation [37, 38, 39]. Second, due to using the claim database, the diagnosis of AMI had the risk of misclassification. We minimized this possibility by defining the diagnosis of AMI and combing examinations for AMI based on the guidelines. Third, the study examined the change in the number of cardiologists, which could be a time-dependent confounding factor. We did not take this point into account; therefore, it is unclear whether the increased proportion of pPCIs in the LM group was influenced by the number of cardiologists.

### Meaning of the study: possible explanations and implications for clinicians and policymakers

This study has shown regional disparities in the proportion of PCIs for AMI patients. Trajectory analysis revealed that the proportion of PCIs was stable over time in three groups and increased yearly in one group. Policymakers should consider such regional differences in the trajectory of this proportion to plan a medical system for AMI practice. This might help to assign the right medical resources in each region equally.

### Unanswered questions and future research

Our study primarily assessed regional disparities in the proportion of PCIs that were related to process indicators. Therefore, further research is required to examine the association between the proportion of PCIs and patient outcome indicators such as cardiovascular death.

## CONCLUSION

The trajectory of the proportion of PCIs for AMI patients identified groups of SMAs. Among the four groups, the LM group showed an increasing trend in the proportion of PCIs, whereas the three other groups showed relatively stable trends. The LM group revealed that the number of cardiologists had increased over time in its regions.

## Data Availability

Data cannot be shared for ethical/privacy reasons.

## iv. Acknowledgments

The current study was funded by JPMH21IA1005 and JPMH21FA1012 of the Ministry of Health, Labour and Welfare (MHLW), and ISHIZUE 2022 of Kyoto University to Yuichi Imanaka. The funders had not play in the study design, data collection and analysis, the manuscript preparation.

