## Supplementay_file for "Regional variations in primary percutaneous coronary intervention for acute myocardial infarction patients: A trajectory analysis using the national claims database in Japan"

**Contents**

Supplementary Figure 1. The population distribution in each group

Supplementary Figure 2. The distribution of the proportion of the population aged 65 years and older by each group

Supplementary Figure 3. The distribution of the total number of medical doctors per 100,000 population in each group

Supplementary Figure 4. The distribution of the number of cardiologists per 100,000 population in each group

Supplementary Figure 5. The distribution of the number of general hospitals per 100,000 population in each group

Supplementary Figure 6. The distribution of the number of general hospital beds per 100,000 population in each group

**Supplementary Figure 1. The population distribution in each group**

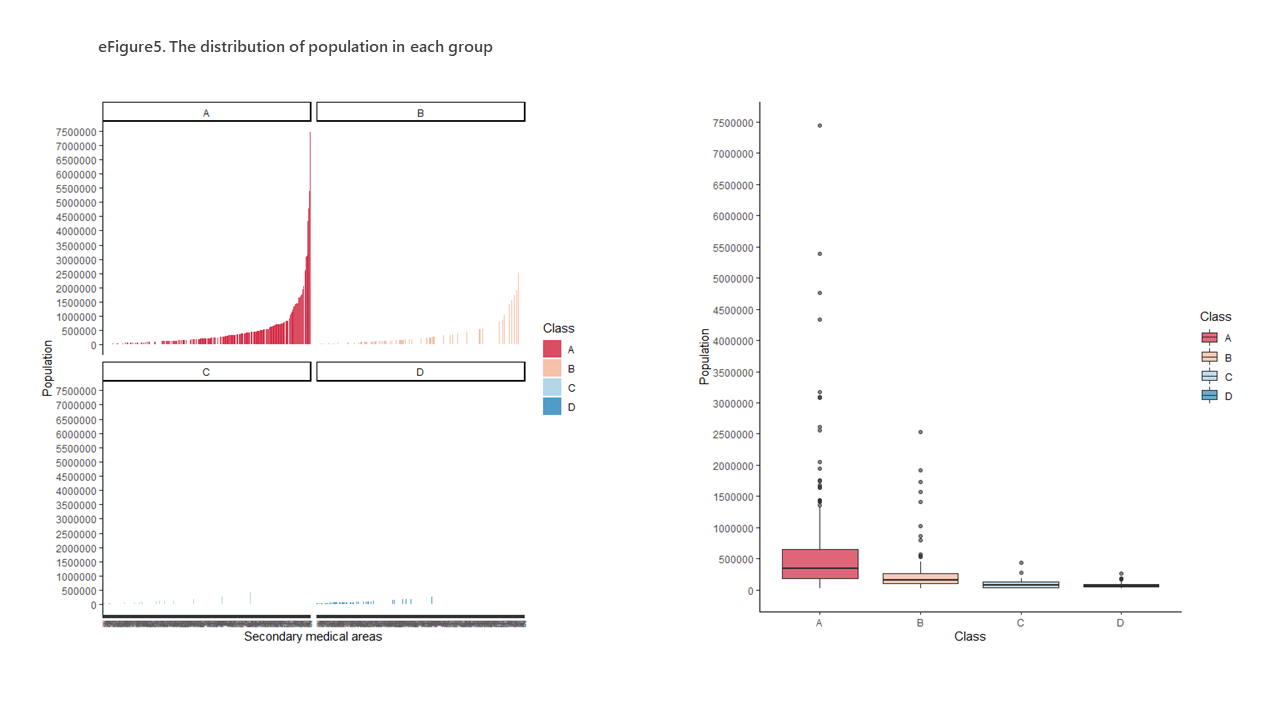

**Supplementary Figure 2. The distribution of the proportion of the population aged 65 years and older by each group**
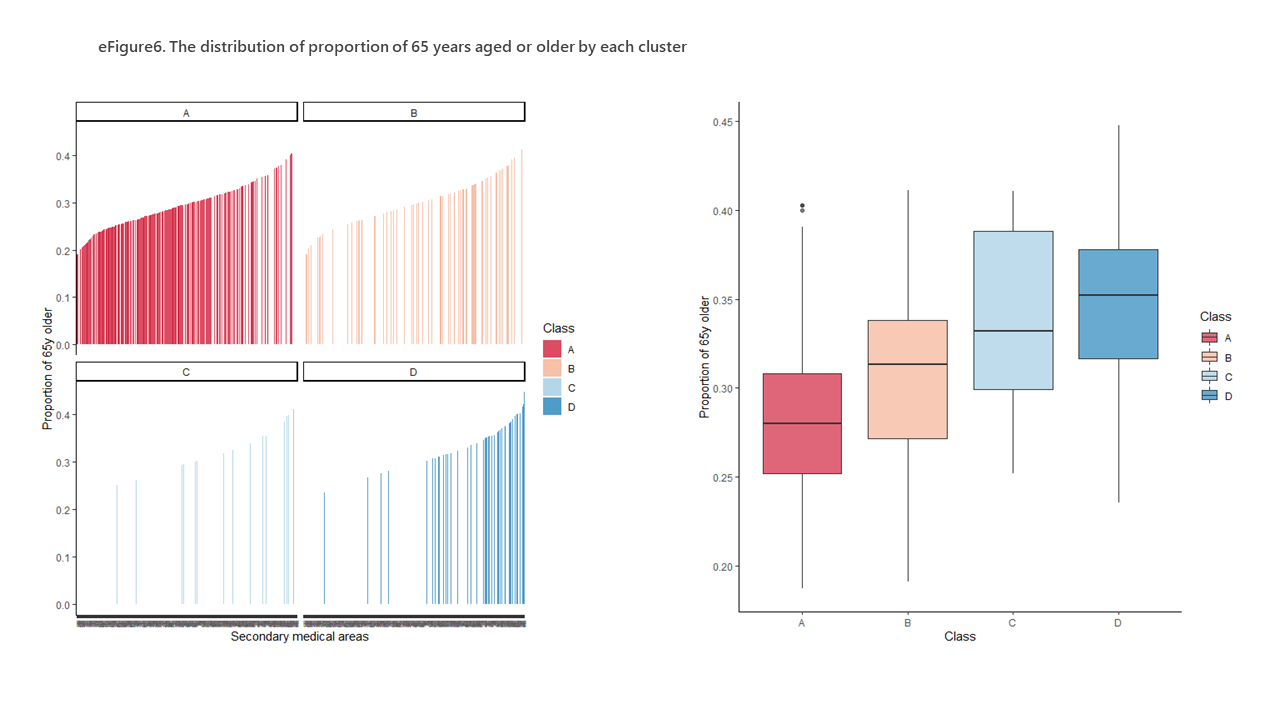

**Supplementary Figure 3. The distribution of the total number of medical doctors per 100,000 population in each group**
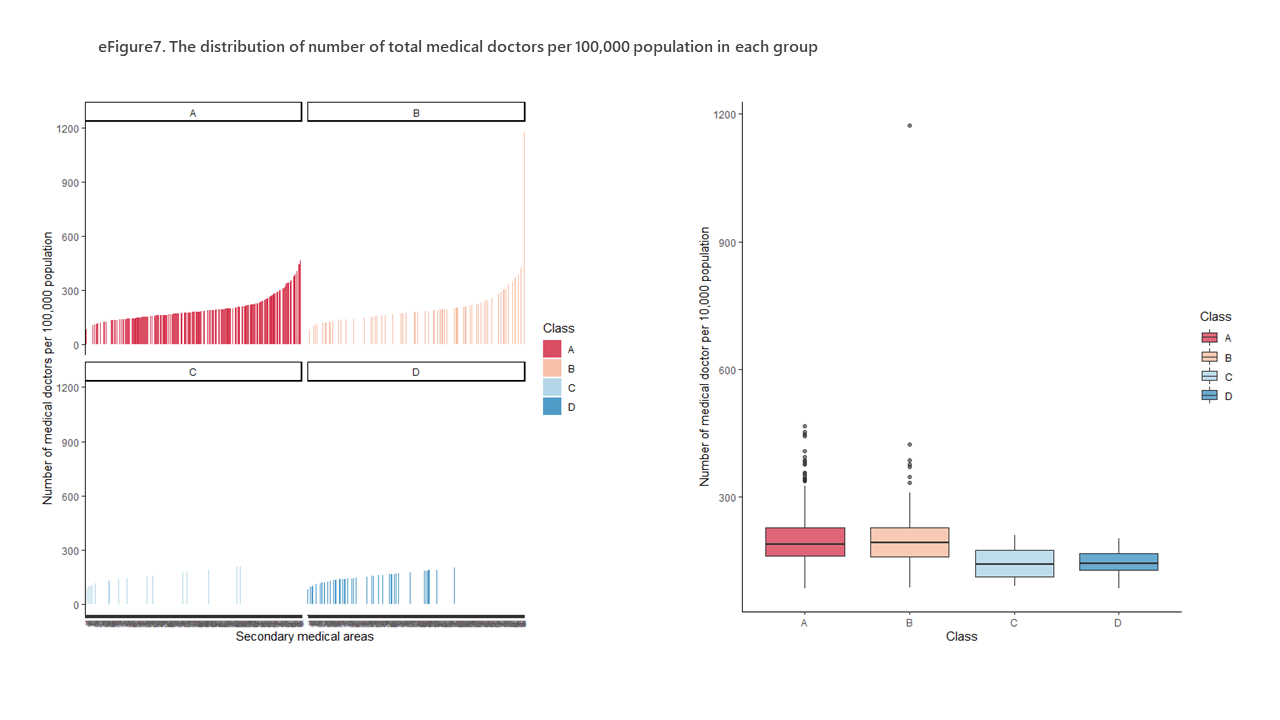

**Supplementary Figure 4. The distribution of the number of cardiologists per 100,000 population in each group**
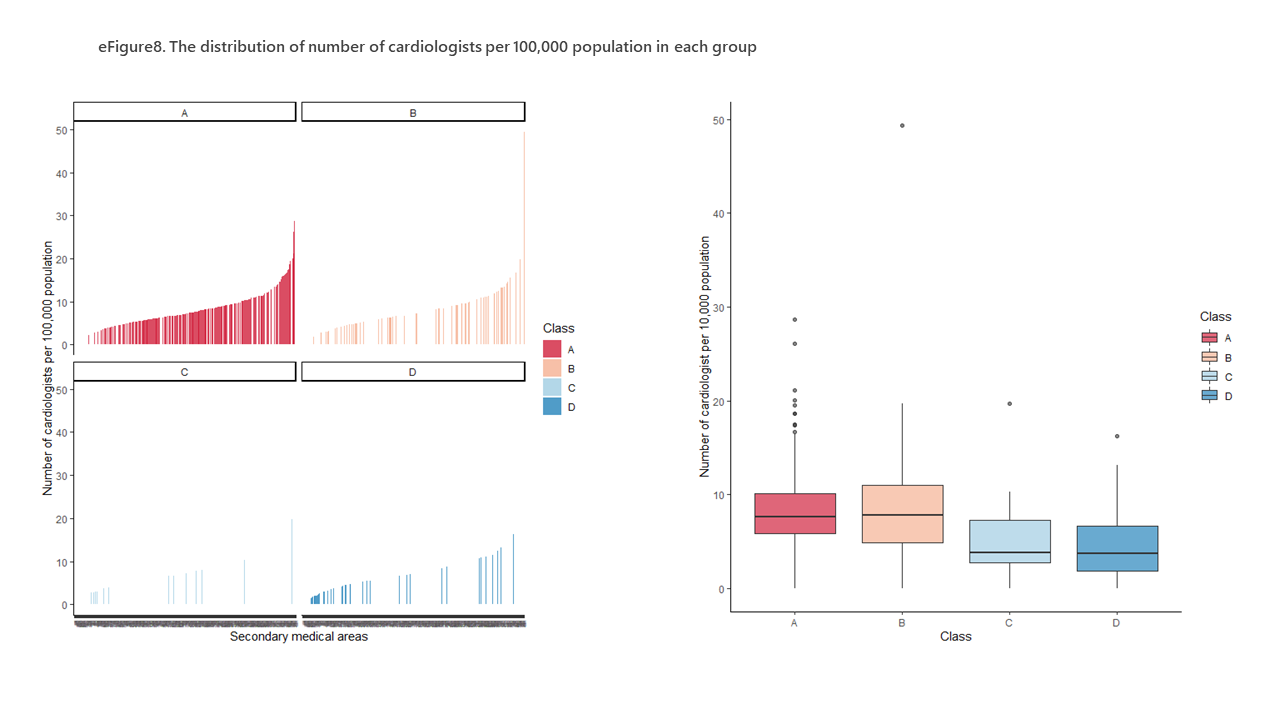

**Supplementary Figure 5. The distribution of the number of general hospitals per 100,000 population in each group**
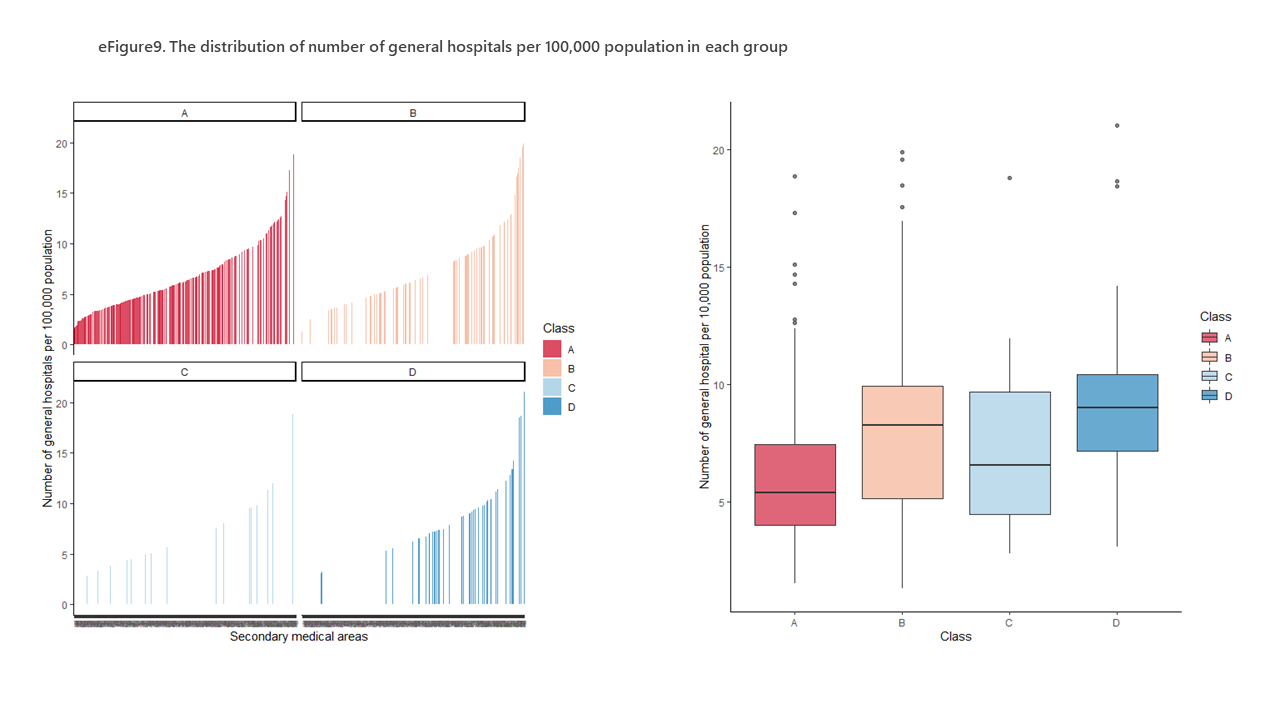

**Supplementary Figure 6. The distribution of the number of general hospital beds per 100,000 population in each group**
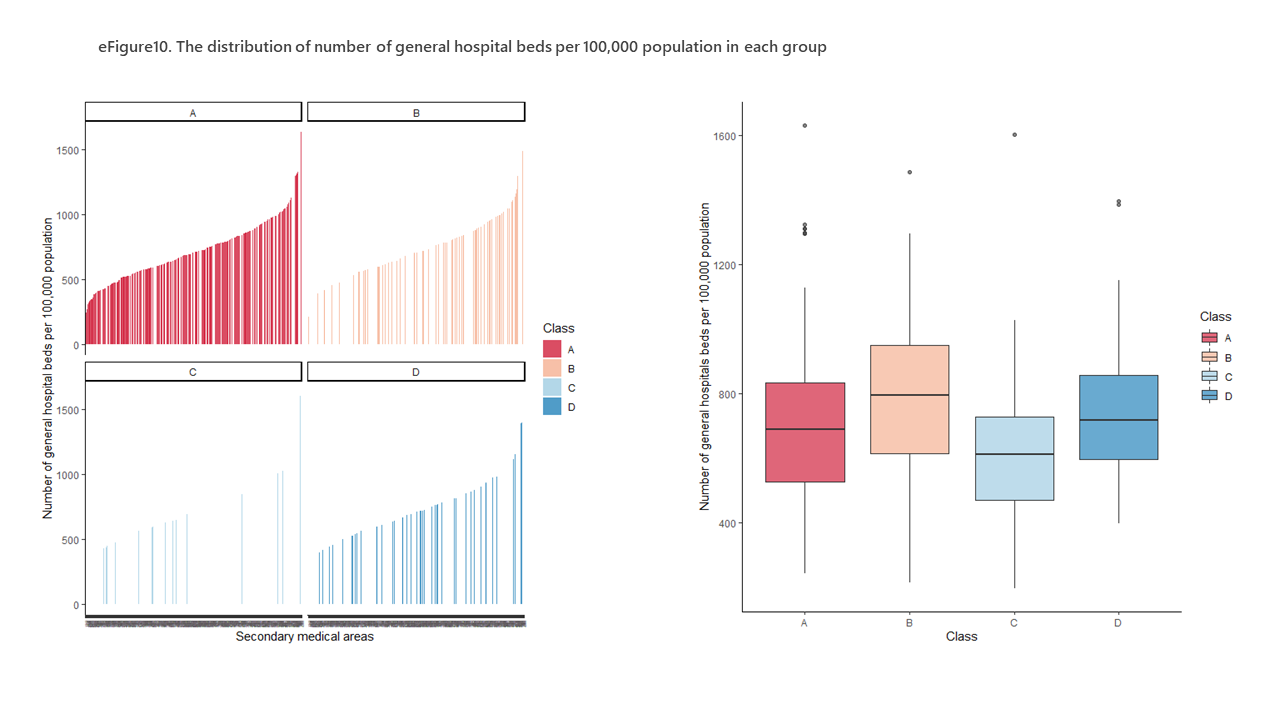
